# Machine learning to predict early recurrence after oesophageal cancer surgery

**DOI:** 10.1101/19001073

**Authors:** Saqib A Rahman, Robert C Walker, Megan A Lloyd, Ben L Grace, Gijs I van Boxel, Feike Kingma, Jelle P Ruurda, Richard van Hillegersberg, Scott Harris, Simon Parsons, Stuart Mercer, Ewen A Griffiths, J.Robert O’Neill, Richard Turkington, Rebecca C Fitzgerald, Timothy J Underwood, On behalf of the OCCAMS Consortium, the full list of contributors is displayed in acknowledgements

## Abstract

**Objective:** To develop a predictive model for early recurrence after surgery for oesophageal adenocarcinoma using a large multi-national cohort.

**Summary Background Data:** Early cancer recurrence after oesophagectomy is a common problem with an incidence of 20-30% despite the widespread use of neoadjuvant treatment. Quantification of this risk is difficult and existing models perform poorly. Machine learning techniques potentially allow more accurate prognostication and have been applied in this study.

**Methods:** Consecutive patients who underwent oesophagectomy for adenocarcinoma and had neoadjuvant treatment in 6 UK and 1 Dutch oesophago-gastric units were analysed. Using clinical characteristics and post-operative histopathology, models were generated using elastic net regression (ELR) and the machine learning methods random forest (RF) and XG boost (XGB). Finally, a combined (Ensemble) model of these was generated. The relative importance of factors to outcome was calculated as a percentage contribution to the model.

**Results:** In total 812 patients were included. The recurrence rate at less than 1 year was 29.1%. All of the models demonstrated good discrimination. Internally validated AUCs were similar, with the Ensemble model performing best (ELR=0.785, RF=0.789, XGB=0.794, Ensemble=0.806). Performance was similar when using internal-external validation (validation across sites, Ensemble AUC=0.804). In the final model the most important variables were number of positive lymph nodes (25.7%) and vascular invasion (16.9%).

**Conclusions:** The derived model using machine learning approaches and an international dataset provided excellent performance in quantifying the risk of early recurrence after surgery and will be useful in prognostication for clinicians and patients.

**DRAFT VISUAL ABSTRACT:** 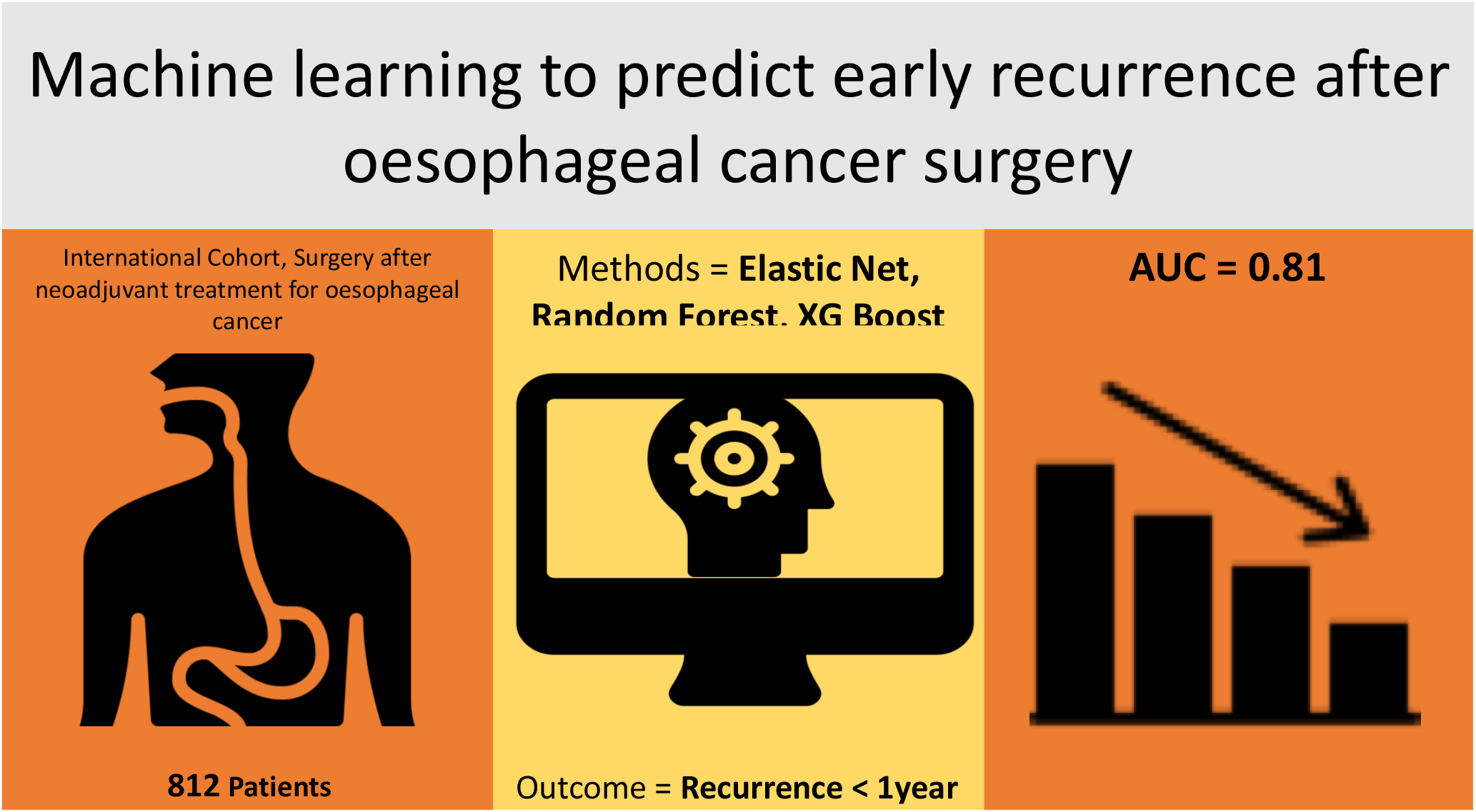

Icons taken from www.flaticon.com, made by ‘Freepik’, ‘smashicons’, and ‘prettycons’. Reproduced under creative commons attribution license

**MINI-ABSTRACT:** Early recurrence after surgery for adenocarcinoma of the oesophagus is common. We derived a risk prediction model using modern machine learning methods that accurately predicts risk of early recurrence using post-operative pathology

## INTRODUCTION

Oesophageal adenocarcinoma carries a poor prognosis. Of the <40% of patients who are candidates for curative treatment^1^, the 5-year survival rate remains approximately 25-50% in randomised trials^2–4^ and rarely in excess of 50% in case series.

Early recurrence (less than 1 year) after surgery is a feared outcome with rates of 20-30%^3–5^ frequently reported, despite the increasing uptake of neoadjuvant chemotherapy (NACT) and chemoradiotherapy (NACRT). This is particularly concerning because recovery from oesophagectomy is often long and the risk of major complications (Clavien-Dindo III-V) is as much as 31.1%^6^. Many patients have not recovered from their primary cancer treatment when they experience cancer recurrence.

In an ideal setting prediction of early recurrence before embarking on a multimodal surgical pathway would provide the most useful information for patients and clinicians. However, staging information correlates poorly between pre- and post-operative settings^7^, and genomic information is not yet able to predict outcome. Even the most robust preoperative models for prediction have an average performance at best^8^. In contrast, postoperative information, although not able to influence surgical treatment decisions, is more prognostic and potentially informative to patients. It may also be helpful in decisions on the merits of adjuvant therapy, further refining the “high risk” group of patients where novel adjuvant treatments are currently being considered.

Naïve logistic regression (LR) has been the dominant approach to binary outcome prediction in clinical medicine for decades. Adoption of modern modified regression and ‘machine learning’ (ML) techniques has been limited, in part due to concerns over computational complexity and reliability. However, an increasing body of evidence demonstrates that they outperform traditional techniques in predictive performance^9,10^, although this is debated^11^. In part, the appeal of these approaches lies in their ability to model complex non-linear relationships which are common in cancer data, and which are challenging to model effectively with logistic/linear approaches. The increasing accessibility of software design now also allows the relatively straightforward deployment of these ‘black-box’ techniques.

Our group has previously published a multicentre UK cohort study which assessed survival according to Mandard Tumour Regression Grade (TRG)^12^. This study included patients who had undergone oesophagectomy for adenocarcinoma of the oesophagus or gastro-oesophageal junction (GOJ) preceded by NACT as part of the Oesophageal Cancer Clinical and Molecular Stratification (OCCAMS) consortium. A clinically meaningful response to NACT was limited to TRG 1-2 only, which represented ∼15% of patients. In the current study we set out to use this database, supplemented with an international cohort from the Netherlands, and machine learning techniques to develop and validate a clinically useful predictive model for early recurrence in oesophageal adenocarcinoma.

## METHODS

### Ethics

The OCCAMS consortium is a UK-wide multicentre consortium to facilitate clinical and molecular stratification of oesophagogastric cancer with ethical approval for biological sample collection and analysis in conjunction with detailed clinical annotation (Research Ethics Committee number: 10/H0305/1). Data collection and participation in research was approved by Institutional ethics committees at each OCCAMS site and UMC Utrecht.

### Source of Data

Data was sourced from 6 tertiary oesophago-gastric centres in the UK, as previously described^12^. Briefly, the records of consecutive patients from each centre between 2000 and 2013 who underwent a planned curative oesophagectomy for adenocarcinoma and also received NACT (platinum-based triplet or cisplatin and 5-Fluorouracil) were reviewed and collated. Clinical, pathological, recurrence and survival data were recorded. Data from one of the original centres was incomplete to the extent that modelling could not take place and was excluded *a priori*. In order to include NACRT as a factor in the model further patients were identified from University Hospitals Southampton (UHS) and University Medical Centre Utrecht (UMCU), where CROSS type NACRT^4^ has been standard of care for oesophageal adenocarcinoma for a number of years. Patients who were deemed irresectable at the time of surgery or who had metastatic disease on the postoperative histology (i.e. pM1) were excluded from analysis.

The primary outcome measure was early recurrence, defined as confirmed local, regional or distant recurrence at less than 1 year from the date of surgery^5,8,13^. Missing Data was treated as being missing completely at random and handled by list wise deletion. Modelling was based on a complete case analysis.

### Predictor Characteristics

Univariate statistics were calculated using Mann Whitney U and Chi-Square test for non-parametric data. The predictive models were generated on the whole dataset (n=812). All available variables were included in the analysis. The circumferential resection margin (CRM) was considered to be involved (and hence R1) in line with Royal College of Pathologists guidelines (i.e. CRM <1mm is positive)^14^. Tumour grade and TRG^15^ were assessed by dedicated gastrointestinal histopathologists who were blinded to clinical data. TRG was considered as responder (TRG1-2) vs non-responder (TRG 3-5) in line with our previous publication using this dataset^12^. To increase the yield of information from lymph node data, both the number of positive lymph nodes and total lymph node harvest were considered as absolute number. For the regression model, linearity was assumed for continuous variables. Explicitly, the variables used to predict outcome were; gender, age, location of tumour, type of neoadjuvant therapy, response to neoadjuvant therapy (TRG), ypT, Vascular invasion, completeness of resection, grade of differentiation, number of positive lymph nodes and total number of lymph nodes examined.

### Model Building and Validation

We elected to use elastic-net regularized logistic regression (ELR)^16^ along with two machine learning techniques; Random Forest (RF)^17^ and Extreme Gradient Boosting (XGboost, XGB)^18^. ELR applies a combination of the ‘ridge’ and ‘lasso’ penalties^19,20^ with the benefits of both (partly minimisation of overfitting and variable selection). RF combines a specified number of decision trees (typically around 1000) created on random subsets of the dataset and is probably the most widely used machine learning approach in medical literature. XGB attempts to improve sequentially by generating models to explain where the original model fails and then repeating this process (typically around 1000 times), while simultaneously applying regularisation to minimise overfitting. Having generated individual models, we then combined them to generate overall predictions^21^, an approach which theoretically is particularly beneficial when using diverse model types (such as those described above) that capture different elements of patients’ risk profiles.

For ELR, the optimal alpha and lambda hyperparameters (penalty severities) were selected by grid-search using 10-fold cross validation with 5 repeats during model generation and ‘log-loss’ as the metric for optimisation. The RF model was derived from 1000 decision trees and hyperparameter tuning was conducted in a similar fashion (for number of variables per tree, split rule and minimum node size). The XGB model was again derived by cross validation of hyperparameters (number of optimisation rounds, maximum tree depth, minimum weight in each child node, minimum loss reduction (gamma), regularization penalty (eta) and subsampling for regularization). Full details of hyperparameter tuning is given in the supplementary materials (S7). These three models were then combined to generate the final (ensemble) model by generating a linear blend of predicted probabilities using logistic regression.

Discrimination of the models was assessed using the area under the receiver operator characteristic (ROC) curve (AUC). In the context of this paper, If two random patients were selected, one with a recurrence of cancer at less than 1 year and one disease free at 1 year, the AUC is equivalent to the probability the model will score the patient with recurrence higher than the patient without. Internal validation was performed using 0.632 bootstrapping, with 1000 resampled datasets. Bootstrapping was preferred for internal validation over splitting the cohort into derivation and validation sets, as this has been shown to reduce bias and improve overall model performance, particularly with moderate datasets^22–24^.Calibration was assessed visually and formally with the Hosmer-Lemeshow Test. As our dataset contains multiple centres with small numbers of patients, we also opted for an internal-external validation procedure, as advocated by Steyerberg and Harrell^25^. This entails generating models on all centres apart from one and validating the model on the remaining centre. This process is then repeated leaving each centre out sequentially and an average calculated.

Unadjusted tree models (such as RF, which is included in the Ensemble) and other maximum margin methods typically calibrate poorly as a consequence of their methodology, with predicted probabilities biased towards the centre. To allow meaningful interpretation of probability, Isotonic regression was used to scale probabilities on the final model, as has been previously described^26,27^.

In contrast to logistic regression, assessing global variable importance is challenging using machine learning techniques and to an extent they are ‘black-boxes’. As coefficients, as would be seen in a logistic regression, are not used an alternative method is required. We used the ‘VarImp’ function of the caret R package, where ROC curves are generated for the outcome for each individual predictor and the contribution to the global ROC curve calculated as a percentage. Due to the nature of higher-order interactions present in the model, variable importance in individual predictions must be calculated independently. We calculated the average marginal contribution of each variable (change from the mean prediction i.e. the Shapley value^28^) for individual predictions. A similar approach was used by Nanayakkara et al. for analysing in-hospital mortality following cardiac arrest^29^.

Data analysis was conducted using R (Version 3.5.3, The R Foundation for Statistical Computing). Models were trained using the ‘caret’^30^ and ‘caretEnsemble’^31^ packages. Individual variable importance was calculated using ‘iml’^32^. All are available at https://CRAN.R-project.org/. Full R code to train the models as described is given in the supplementary materials (S7), along with a list of packages used.

The calibrated final model was designed using R Shiny^33^ and is available freely at: https://uoscancer.shinyapps.io/EROC/. No data entered into the model is collected or stored.

## RESULTS

A total of 812 patients from 7 centres were included in model training. A consort diagram detailing patient numbers and the final sample size is presented in Figure 1.

**Figure 1:**
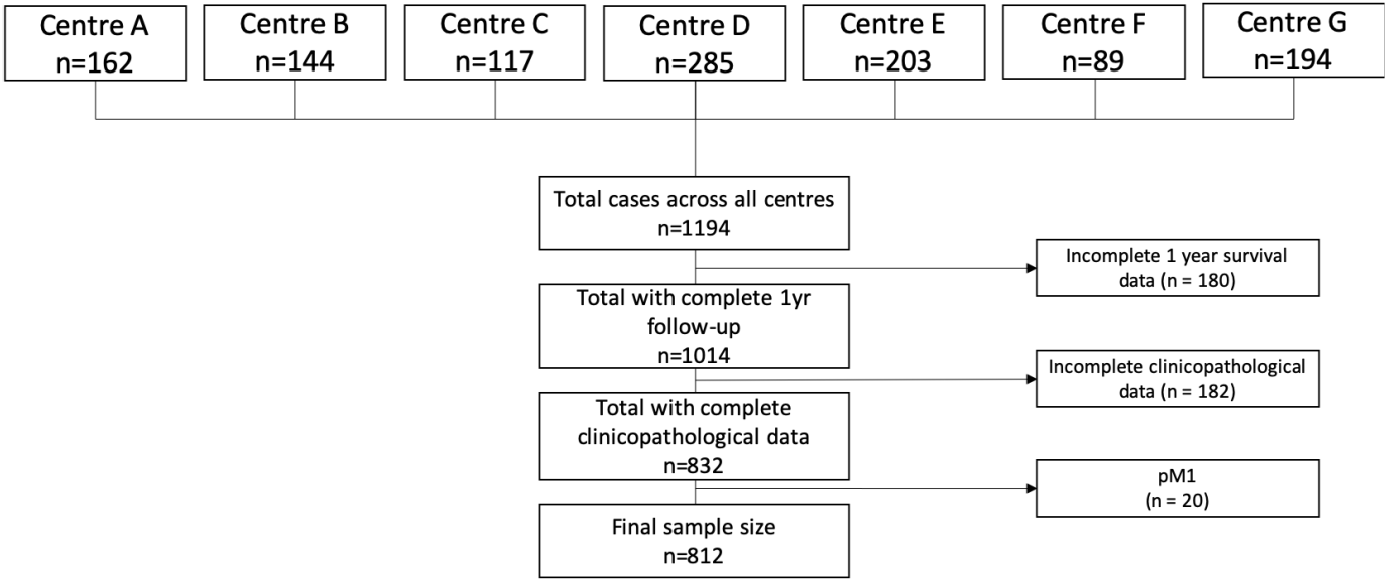
Consort Diagram.

Most patients were male (84.6%), with a median age of 64 years. The majority of tumours were at the GOJ (55.5%), with a high proportion of locally advanced (ypT3-4 – 66.8%) and node positive disease (61.0%). First recurrence of cancer within 1 year of surgery was identified in 236 patients (29.1%). The early recurrence group were significantly less likely to have responded to neoadjuvant treatment (8.5% vs 21.7%), and had worse ypT, ypN, Vascular invasion, R1 resection rate and grade of differentiation (all p<0.001). Detailed group clinicopathological data is shown in Table 1.

**Table 1:** Cohort and Group Clinicopathological Data.

| Characteristic |  | All Patients<br>(n = 812) | No Early Recurrence<br>(n = 576) | Early Recurrence<br>(n = 236) | P§ |
| --- | --- | --- | --- | --- | --- |
| Gender | Male | 687 (84.6) | 487 (84.5) | 200 (84.7) | 0.944 |
|  | Female | 125 (15.4) | 89 (15.5) | 36 (15.3) |  |
| Age |  | 64 (28-83) | 63.9 (28-81) | 64.1 (38-83) | 0.855¶ |
| Site | Oesophagus | 361 (44.5) | 250 (43.4) | 111 (47) | 0.352 |
|  | GOJ | 451 (55.5) | 326 (56.6) | 125 (53) |  |
| TRG | 1-2 | 145 (17.9) | 125 (21.7) | 20 (8.5) | <0.001* |
|  | 3-5 | 667 (82.1) | 451 (78.3) | 216 (91.5) |  |
| ypT | 0 | 33 (2.6) | 28 (4.9) | 5 (2.1) | <0.001* |
|  | 1 | 96 (11.8) | 87 (15.1) | 9 (3.8) |  |
|  | 2 | 141 (17.4) | 125 (21.7) | 16 (6.8) |  |
|  | 3 | 495 (61.0) | 320 (55.6) | 175 (74.2) |  |
|  | 4 | 47 (5.8) | 16 (2.8) | 31 (13.1) |  |
| Positive LN (n) |  | 1 (0-41) | 0.5 (0-30) | 4 (0-41) | <0.001*¶ |
| ypN >0 |  | 495 (61.0) | 288 (50) | 207 (87.7) | <0.001* |
| Total LN (n) |  | 24 (0-75) | 24 (0-75) | 23 (6-61) | 0.805¶ |
| Total LN >15 |  | 688 (84.7) | 481 (83.5) | 207 (87.7) | 0.134 |
| Vascular Invasion |  | 372 (45.8) | 202 (35.1) | 170 (72) | <0.001* |
| R1 Resection |  | 236 (29.1) | 118 (20.5) | 113 (47.9) | <0.001* |
| Grade | Well | 63 (7.8) | 55 (9.5) | 8 (3.4) | <0.001* |
|  | Moderate | 300 (36.9) | 233 (40.5) | 67 (28.4) |  |
|  | Poor/Anaplastic | 449 (55.3) | 288 (50) | 161 (68.2) |  |
| Neoadjuvant Treatment | NACT | 657 (80.9) | 476 (82.6) | 181 (76.7) | 0.061 |
|  | NACRT | 155 (19.1) | 100 (17.4) | 55 (23.3) |  |

Data presented as absolute number (%) and median (range), \*p<0.05. § $\chi^2$ test, except ¶ Mann-Whitney U test

### Model performance (discrimination)

Discrimination was assessed in the training set, internally (via bootstrapping) and internally-externally (across centres). ROC curves of the internal validation of each model are shown in Figure 2. All models demonstrated excellent discrimination on the training set (apparent discrimination), with the Random Forest Model performing the best (AUC 0.98), followed by the Ensemble model (0.90), XGB (0.85) and ELR (0.81). On internal validation, the Ensemble model had the best performance (AUC 0.81) and the ELR the worst (AUC 0.79). Overall discrimination for each model is summarised in table 2.

**Table 2:** Model Discrimination AUC (95% CI)

| Model | Apparent | Internal Validation | Internal-External Validation |
| --- | --- | --- | --- |
| Elastic Net | 0.805 (0.772-0.838) | 0.785 (0.783-0.786) | 0.798 (0.713-0.883) |
| Random Forest | 0.980 (0.972-0.987) | 0.789 (0.787-0.790) | 0.805 (0.721-0.889) |
| XG Boost | 0.849 (0.822-0.877) | 0.794 (0.792-0.796) | 0.800 (0.716-0.883) |
| Ensemble | 0.902 (0.881-0.992) | 0.806 (0.799-0.812) | 0.804 (0.721-0.887) |

**Figure 2:**
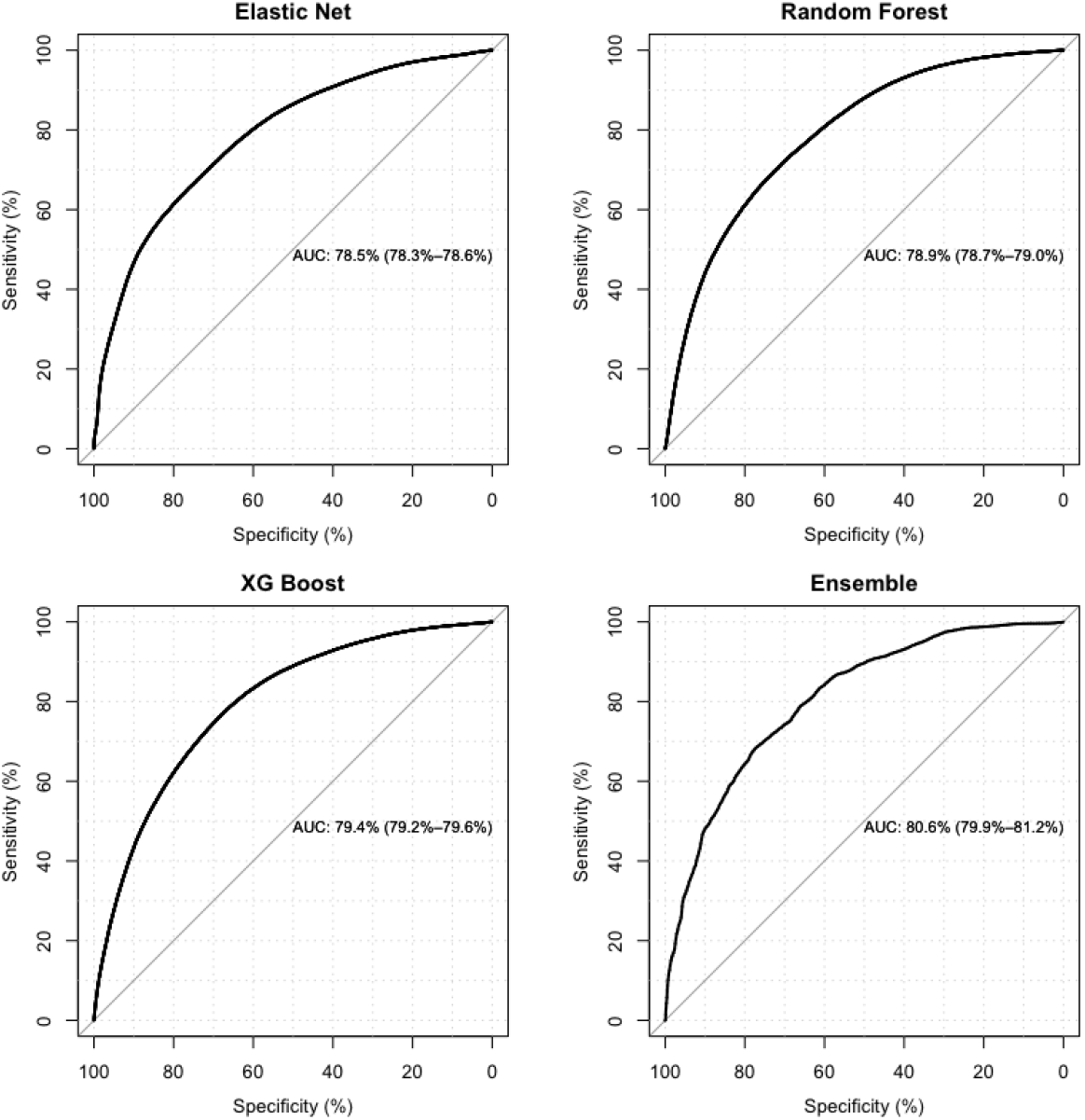
Model discrimination on internal validation by 0.632 bootstrap.

### Model performance (calibration)

Calibration on the training set was visually best in the ELR, and worst in the RF and Ensemble models (supplementary materials). This was corroborated by the Hosmer Lemeshow test (p value ELR=0.806, RF=<0.001, XGB=0.030, Ensemble=<0.001). Probabilities generated by the final model were scaled using isotonic regression. Calibration before and after scaling is shown in Figure 3 (shaded area represents two standard errors, calibration tables in supplementary materials). The Hosmer Lemeshow test before scaling gives a Chi2 of 38.0 and p<0.001, and after gives Chi2 of 4.5 and p=0.806. Similarly, the Brier score (a measure of overall model performance) also improves from 0.119 to 0.114.

**Figure 3:**
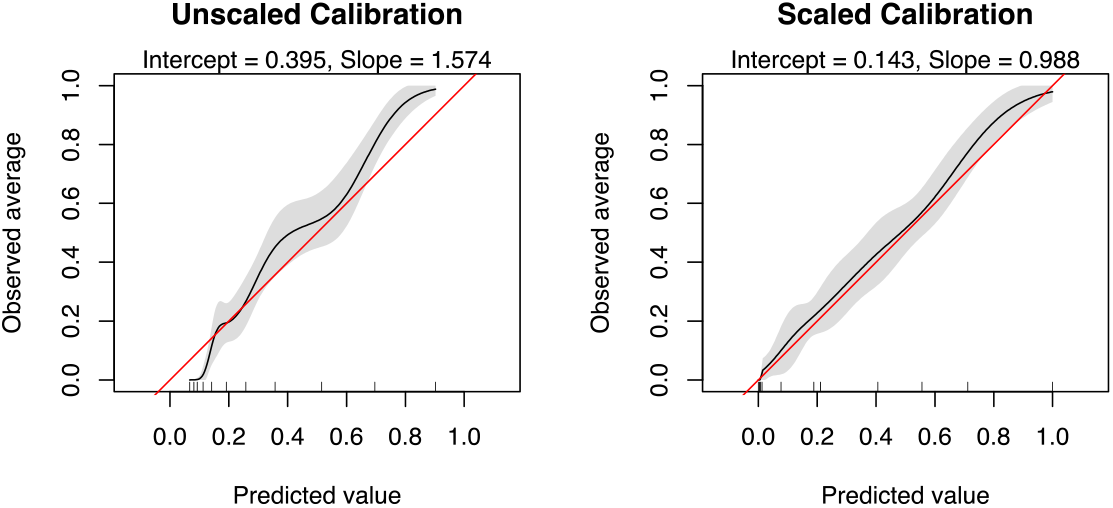
Final model calibration before and after adjustment.

### Variable Importance

Coefficients and odds-ratios cannot be generated for these models. We therefore computed variable importance as a percentage contribution to the model. The results are displayed in Table 3.

**Table 3:** Variable Importance.

| Predictor Variable | Elastic Net (%) | Random Forest (%) | XG Boost (%) | Final Model (%) |
| --- | --- | --- | --- | --- |
| Gender | 0.0 | 1.1 | 1.2 | 0.8 |
| Age | 0.3 | 18.2 | 10.2 | 9.6 |
| Site of Tumour | 9.4 | 2.6 | 4.8 | 5.6 |
| Response to Neoadjuvant Treatment | 0.0 | 0.0 | 0.0 | 0.0 |
| ypT | 11.2 | 9.2 | 7.4 | 9.2 |
| Number of Positive Lymph Nodes | 3.6 | 30.8 | 40.9 | 25.7 |
| Nodes Examined | 0.4 | 16.7 | 7.0 | 8.0 |
| Vascular Invasion | 26.8 | 10.5 | 13.6 | 16.9 |
| Completeness of Resection (R0/R1) | 15.9 | 5.2 | 6.1 | 8.9 |
| Grade | 7.0 | 3.2 | 2.1 | 4.0 |
| Neoadjuvant Treatment (CT/CRT) | 25.4 | 2.5 | 6.8 | 11.4 |

**Table 4:**
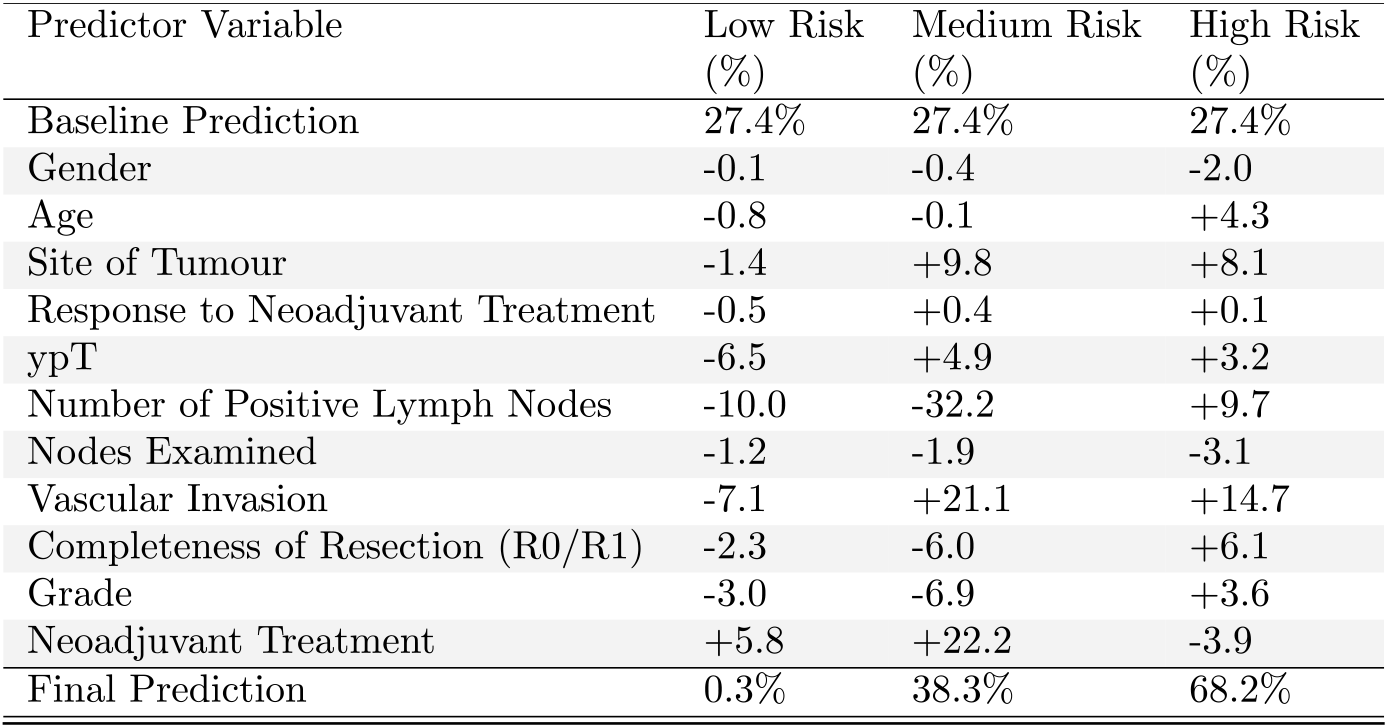
Patient Examples using final model. This table illustrates the percentage contribution of each variable in each setting, in terms of a change from the baseline prediction, 27.4%. A calculator for this is provided with the online model.

Overall the most influential predictor variable is number of positive lymph nodes (25.7%), followed by Vascular Invasion (16.9%). There is considerable variability in importance across models. For example, Age contributes 0.3% to the ELR model, 18.2% to the RF model, 10.2% to the XGB model and 9.6% to the Final model.

It is important to restate that the relationships between the variables and outcome are non-linear and their importance varies considerably according to other variables due to higher order interactions. As an example, even though lymph node status is the most influential marker overall, there are combinations of other variables that would make other variables most important in individual patients. To illustrate this and demonstrate how variables interact, three example patients are considered below. The technique used measures the change in the prediction from the mean prediction (27.1%) that can be attributed to each predictor variable. This approach (calculation of the Shapley value) originates from cooperative game theory.

#### Example 1: Low Risk Patient (AJCC ypT0N0M0: Stage 1)

*50 year old Male with a GOJ adenocarcinoma who undergoes neoadjuvant chemoradiotherapy. On postoperative pathology he is a responder with ypT0, negative vascular invasion, R0 resection and a well differentiated tumour. He has 0 positive lymph nodes out of 30 sampled*.

#### Example 2: Medium Risk Patient (AJCC ypT3N0M0: Stage 2)

*66 year old Male, with an Oesophageal adenocarcinoma who undergoes neoadjuvant chemoradiotherapy. On postoperative pathology he is a non-responder with ypT3, positive vascular invasion, R0 resection and a moderately differentiated tumour. He has 0 positive lymph nodes out of 30 sampled*.

#### Example 3: High Risk patient (AJCC ypT3N2M0: Stage 3b)

*70 year old Female with an Oesophageal adenocarcinoma who undergoes neoadjuvant chemotherapy. On postoperative pathology she is a non-responder with ypT3, positive vascular invasion, R1 resection, Poor differentiation with 5 positive lymph nodes out of 30 sampled*.

## DISCUSSION

In this study we have derived an easy to use and robust clinical model for predicting the risk of early recurrence after surgery for oesophageal adenocarcinoma. It uses routinely collected clinical and pathological data which should be available for every patient. The final model demonstrated excellent discrimination, and validation techniques supported the generalisability of the approach.

In addition to prognostication, this model may be useful for planning adjuvant therapy. Early recurrence after oesophagectomy, often before recovery from surgery is complete, is a devastating outcome for patients. Targeting existing and emerging treatment combinations in this patient group to prolong time to recurrence or prevent recurrence is vital, however can only happen with accurate predictions of the likelihood of relapse. We have purposefully avoided dichotomization/stratification based on outcome and presented raw probability in preference to this. This will allow full discussions between surgeons/oncologists and patients to take place regarding the benefits of adjuvant therapy and tailored to individual patient’s post-operative recovery and wishes. It may also allow stratification of adjuvant trials based on layered levels of risk.

This cohort exhibited an early recurrence rate of 29.1%, which is similar to previous reports^3–5,8^. There was an R1 resection rate of 29.1%, in line with previously reported data^34,35^ with an RCP definition of CRM positivity (CRM<1mm involved). On univariate analysis all factors expected to correlate with worse prognosis (including ypT, ypN, vascular invasion, R1 resection and grade of differentiation) were significantly worse in those patients who developed an early recurrence. This validates our cohort as a true representation of contemporary practice and a sensible place to begin building more complex models.

Discrimination of the different models was similar, with minimal variability of AUC between models on validation. However, the ensemble model consistently performed the best and is a suitable choice for the final model. The decline in performance from the training set to validation, which was particularly marked in the RF and ensemble models, is a consequence of the tuning process, whereby the optimum values are chosen from a grid of thousands after repeated tests (in this case repeated 10-fold cross validation). In this setting, the apparent performance of the model on the training set is over-estimated and should be disregarded.

There was marked heterogenicity in variable importance between models. This is interesting, particularly in the context of the models performing so similarly overall and supports the idea of combining them to capture different patient information. The most important variables overall were number of positive lymph nodes and vascular invasion, accounting for 42.6% of performance. This is not only biologically sensible, but the subject of several recent publications and ongoing translational work^12,36,37^. Although not available for this study, more detail regarding lymphadenopathy – e.g. downstaging and anatomical location would likely be informative. Firm conclusions regarding variables are difficult considering the nature of the study. However, we would draw attention to two facets of the model. Firstly, TRG was the least influential variable across the board, with an importance of almost 0%. This suggests that in itself TRG adds no information over the other measured variables in predicting early outcomes. This is in keeping with emerging data regarding the genomic disparity between primary tumours and their metastasis (lymph node or distant) and our previous report of the importance of lymph node downstaging to clinical outcome. Secondly, modality of treatment was the third most important determinant of outcome, with NACT conferring an advantage over NACRT. In this cohort, despite having considerably better postoperative pathology after NACRT, the rate of early recurrence was no less, and borderline higher (Supplement 4). This suggests that although there is superior post-operative pathology seen with NACRT, this does not translate to better outcome^38–41^ and hence a ypT3N1R0 after NACT does not have the same meaning as a ypT3N1R0 result after NACRT, at least in the early period after treatment. This is important in postoperative discussions with patients.

Other risk factors for early recurrence including perioperative blood transfusion^42^, complications of surgery^43^ and preoperative staging were not available for this study, but are less discriminatory. Precise neoadjuvant regimens were not available for all patients in this study. It is therefore unclear if these results would be influenced by completion of treatment as prescribed, or indeed any adjuvant therapy given. This seems to have minimal effect on the model and suggests a small margin of effect on outcomes. Combining these factors could potentially increase the performance of our model if incorporated in the future. Ultimately, differential gene expression and mutation^44,45^ may well determine prognostication and treatment pathways^46^, but we are likely years from this being universally available. Until then clinical and histopathological data remains the gold standard.

In that context, gains from mathematical and computer-based techniques are key to precision in delivery of cancer care. Here we have demonstrated several modern approaches that produce viable models. This study uses a dataset which is relatively small and simple in a ML context, and the improvement in performance over a standard LR is small (internal validation AUC 0.781). This is none-the-less important as this improvement is in effect ‘free’. The strengths of this study lie in its multi-centre nature and heterogenicity of the cohort. This approach should maximise the utility of the model on external populations. All the data points used should be collected routinely at the majority of institutions, which should allow uptake without change in practice. The College of American Pathologists (CAP) definition of CRM positivity (i.e. CRM positive if tumour at the resection margin) was derivable for Centre G and performance was preserved in this subgroup if used instead of RCP definition (AUC of 0.813 with model generated on centres A-F (n=650) and validated on centre G (n=162), supplementary materials 5), supporting utility in both settings. We have also focussed on predictive model study design and reporting as suggested by the AJCC^47^ and TRIPOD statements^48^.

The training set was limited to patients undergoing neoadjuvant therapy for adenocarcinoma of the oesophagus. We have made no attempt to apply the model to a chemotherapy naïve population, and it is unlikely to calibrate well in this group due to the differing influence of survival of ‘yp’ compared to ‘p’ staging^49^. It is also unclear if the model would be valid in patients with squamous cell carcinoma and we would advocate an early external validation exercise using this patient group. A formal reassessment/recalibration using the CAP definitions of CRM positivity would also be beneficial. Simulation studies have suggested that 100 – 200 cases (i.e. positives) are required for accurate validation^50^.

Assuming a stable incidence this would require approximately 380 – 760 patients. A further limitation was the significant proportion of the original cases with missing data, which will have introduced a degree of selection bias. Multiple imputation is possible as a means of addressing this, however was felt less appropriate in this study due to the high proportion of missing data being in the outcome measure and the lack of an external validation set.

## Conclusion

This large, multicentre cohort of patients who underwent oesophagectomy has been used to derive an accurate prediction model for early cancer recurrence, with excellent performance on an external cohort. Machine learning techniques represent an attractive proposition for maximising performance of predictive models. The model is presented for use at https://uoscancer.shinyapps.io/EROC/.

## Data Availability

Data referred to in the manuscript was collected in the specified centres as part of local quality assurance and the OCCAMS project and remains property of those institutions

## Acknowledgements

Rogier van der Sluijs, UMC Utrecht, provided advice on predictive model methodology

## SUPPLEMENTARY MATERIALS

### S1 Centre Clinicopathological Information

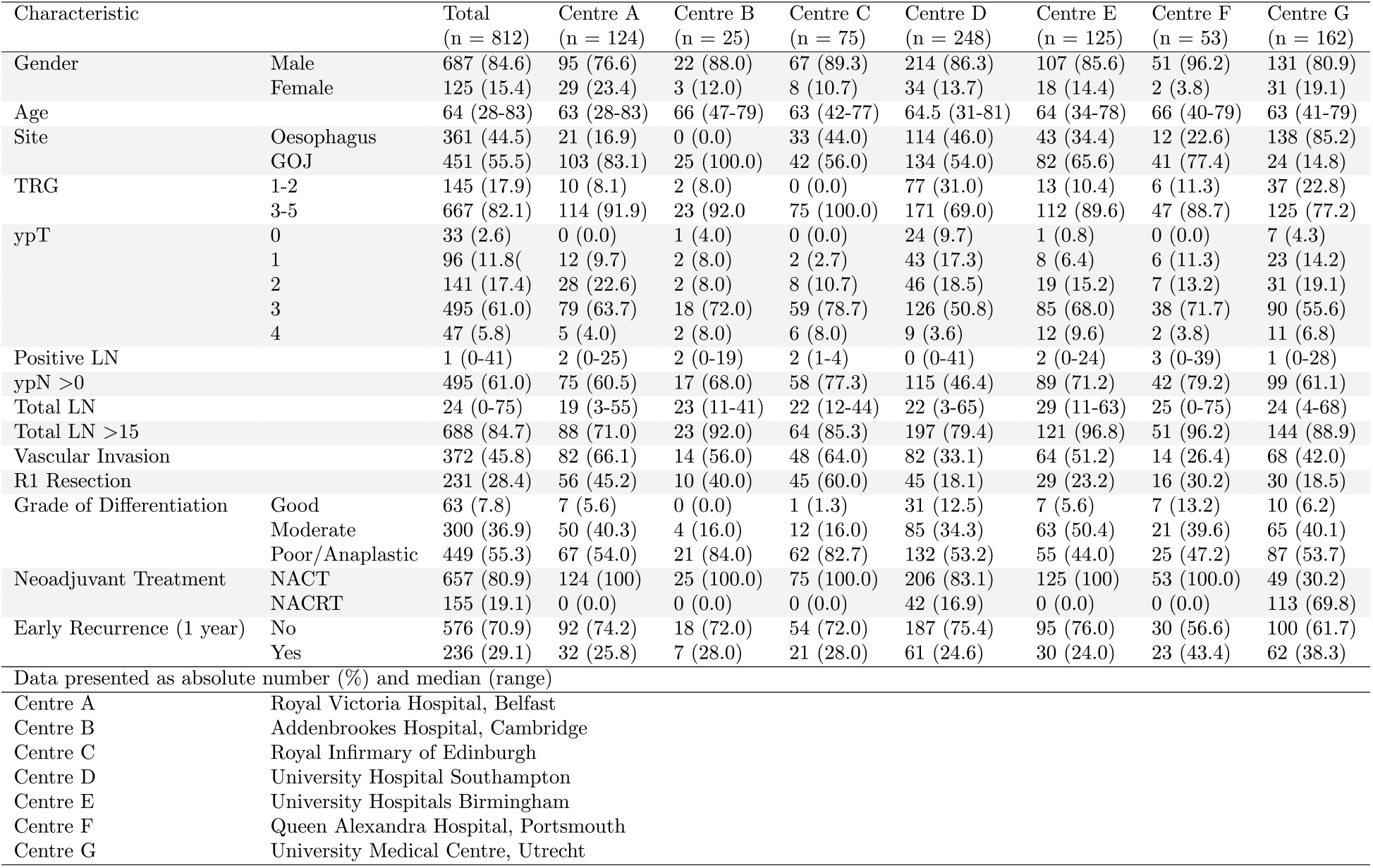

### S2 Model Calibration

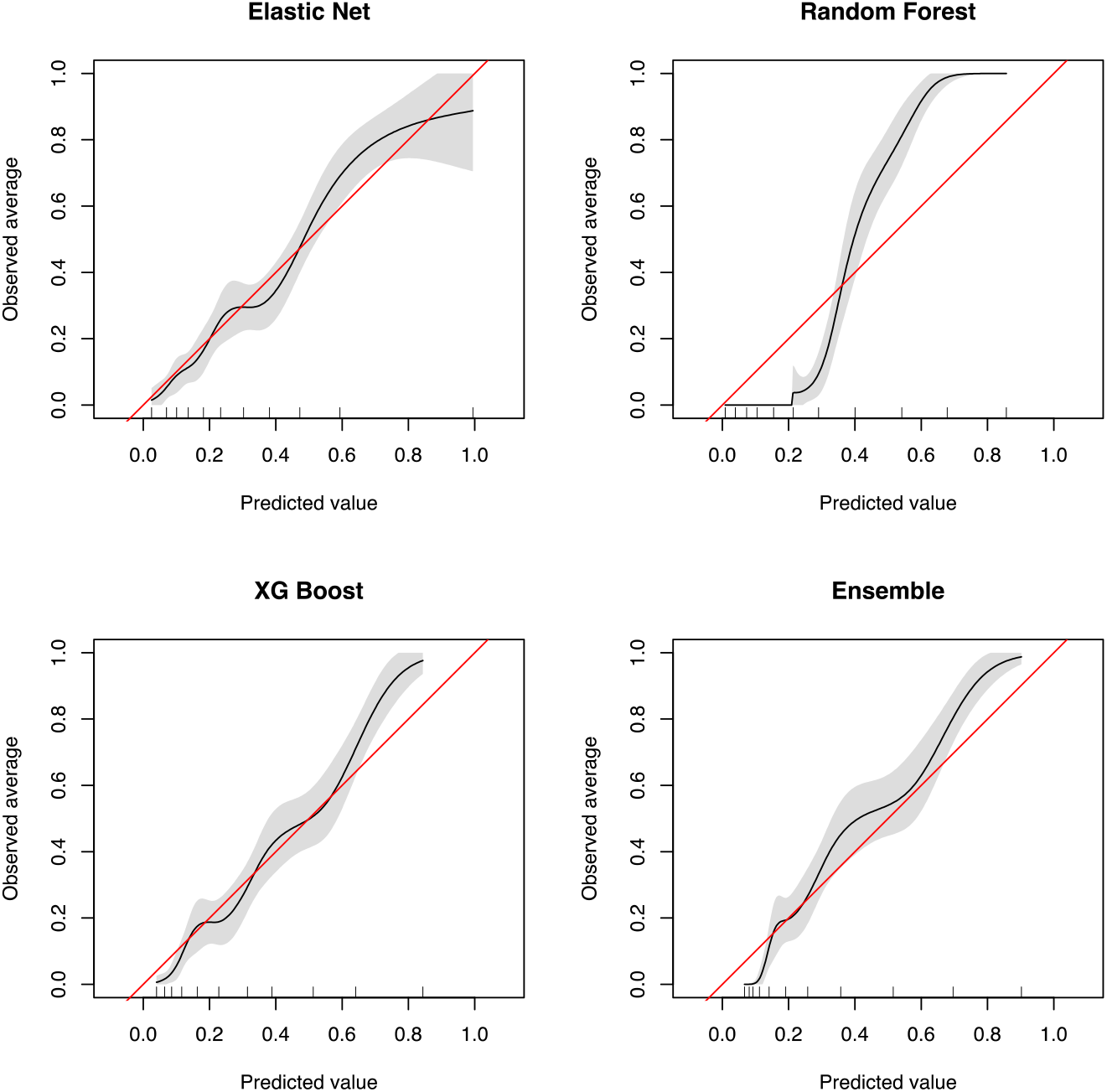

### S3 Calibration of Final model before and after adjustment

| Adjustment | Cases | 0-10% | 10-20% | 20-30% | 30-40% | 40-50% | 50-60% | 60-70% | 70-80% | 80-90% | 90-100% |
| --- | --- | --- | --- | --- | --- | --- | --- | --- | --- | --- | --- |
| Before | Expected | 16.6 | 30.1 | 25.8 | 24.3 | 23.4 | 24.3 | 28.7 | 29.8 | 32.6 | 1.8 |
|  | Observed | 0 | 21 | 26 | 30 | 29 | 23 | 33 | 34 | 38 | 2 |
| After | Expected | 7.5 | 11.5 | 6.8 | 25.2 | 5.5 | 49.5 | 26.5 | 22.7 | 16.2 | 48.5 |
|  | Observed | 7 | 27 | 11 | 7 | 27 | 50 | 30 | 14 | 21 | 42 |

**S4: NACT and NACRT characteristics**

### S4 NACT and NACRT characteristics

| Characteristic |  | NACT<br>(n = 657) | NACRT<br>(n = 155) | P§ |
| --- | --- | --- | --- | --- |
| Gender | Male | 554 (84.3) | 133 (85.8) | 0.711 |
|  | Female | 103 (15.7) | 22 (14.2) |  |
| Age |  | 64 (28-83) | 65 (41-79) | 0.855¶ |
| Site | Oesophagus | 232 (35.3) | 129 (83.2) | <0.001* |
|  | GOJ | 425 (64.7) | 26 (16.8) |  |
| TRG | 1-2 | 91 (13.9) | 54 (34.8) | <0.001* |
|  | 3-5 | 566 (86.1) | 101 (65.2) |  |
| ypT | 0 | 17 (2.6) | 16 (10.3) | <0.001* |
|  | 1 | 71 (10.8) | 25 (16.1) |  |
|  | 2 | 114 (17.4) | 27 (17.4) |  |
|  | 3 | 415 (63.2) | 80 (51.6) |  |
|  | 4 | 40 (6.1) | 7 (4.5) |  |
| Positive LN |  | 1 (0-39) | 1 (0-41) | 0.002*¶ |
| ypN >0 |  | 411 (62.6) | 84 (54.2) | <0.001* |
| Total LN |  | 23 (0-75) | 25 (9-68) | 0.010*¶ |
| Total LN >15 |  | 545 (83) | 143 (92.2) | 0.003* |
| Vascular Invasion |  | 320 (48.7) | 52 (33.5) | <0.001* |
| R1 Resection |  | 218 (33.2) | 13 (8.4) | <0.001* |
| Grade of Differentiation | Well | 47 (7.2) | 16 (10.3) | 0.038* |
|  | Moderate | 233 (35.5) | 67 (43.2) |  |
|  | Poor/Anaplastic | 377 (57.4) | 62 (40) |  |
| Early Recurrence (1 year) | No | 476 (72.5) | 100 (64.5) | 0.061 |
|  | Yes | 181 (27.5) | 55 (35.5) |  |

Data presented as absolute number (%) and median (range), \*p<0.05.
§ $\chi^2$ test, except ¶ Mann–Whitney U test

### S5 Model Discrimination on Centre G Cohort using CAP criteria for CRM

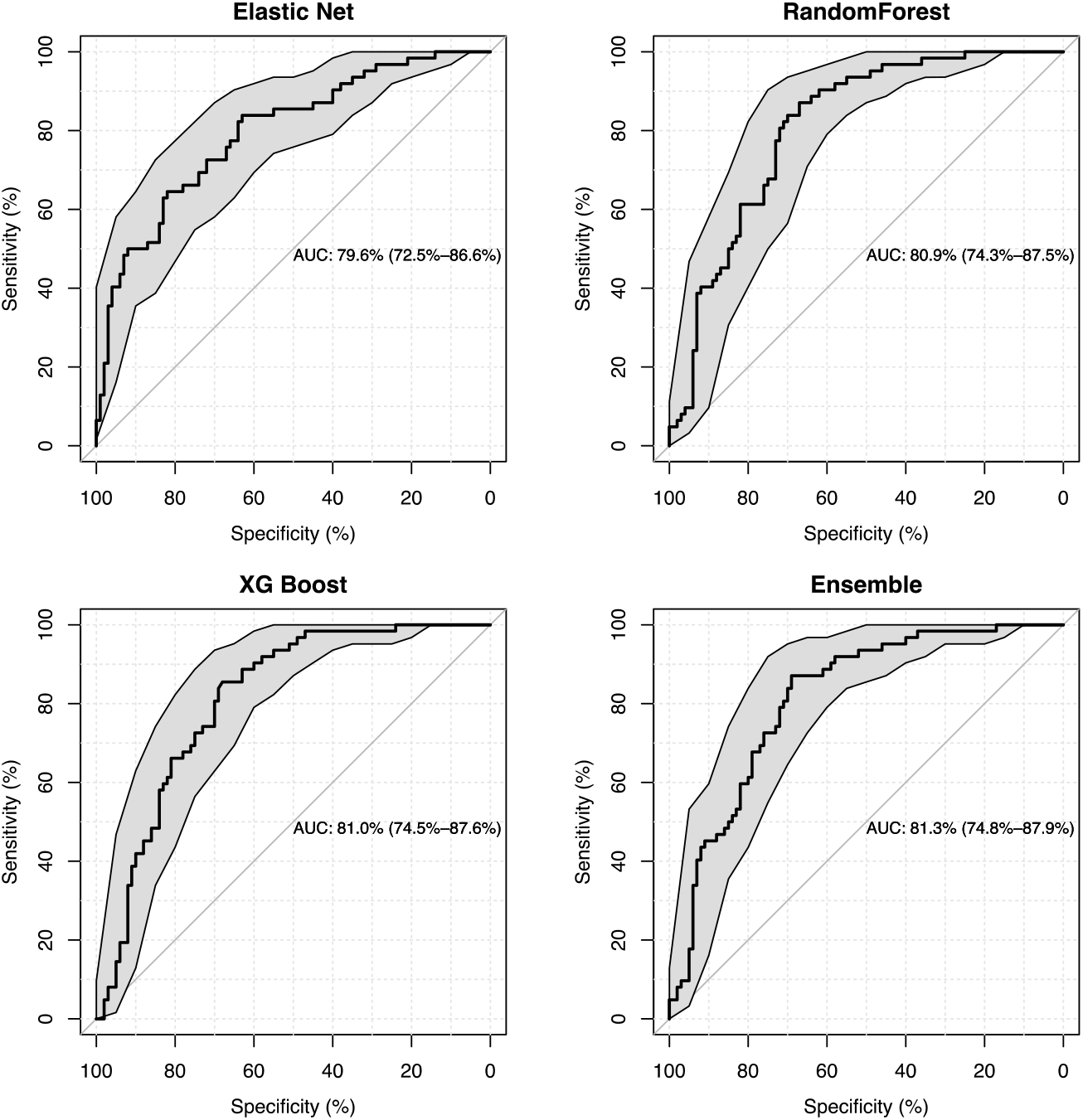

### S6 Internal-External Validation ROC Curves of final model

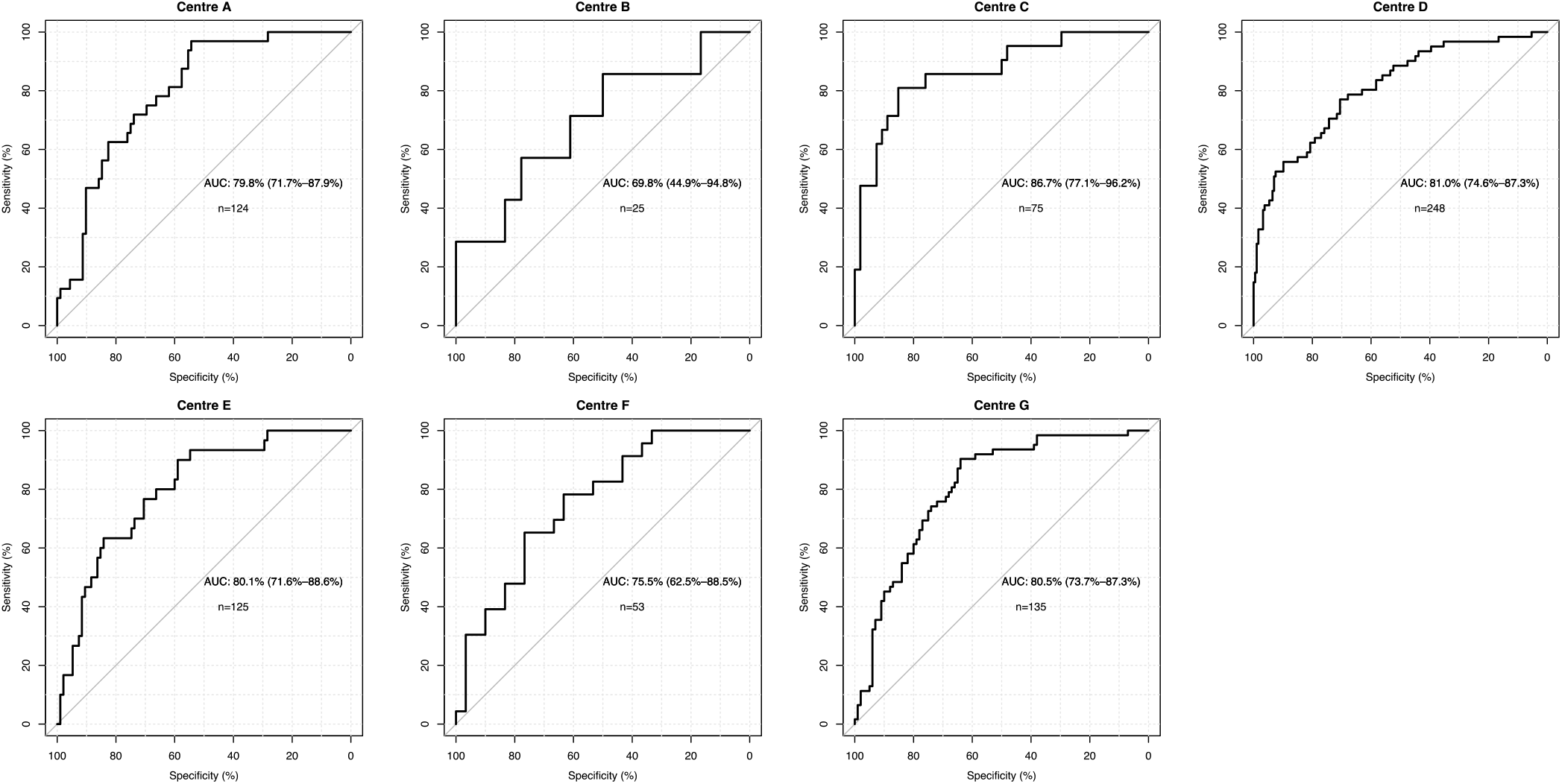

### S7 R-Code

~~~
####The below code will train an elastic net, random forest, XGboost and ensemble models on data
provided, and internally validate them. Descriptions of how to use the model on batches of new data (e.g.
for validation are also provided)
####Saqib A Rahman June 2019.

####Installs required packages
packages <- c(“caret”, “pROC”, “gbm”, “caretEnsemble”, “ResourceSelection”)
if (length(setdiff(packages, rownames(installed.packages()))) > 0) {
 install.packages(setdiff(packages, rownames(installed.packages())))
}

####Loads required packages
library(caret) ###Trains the models
library(pROC) ###Generates receiver operator characteristic curves
library(gbm) ###Generates Calibration plot
library(caretEnsemble)###Combines models
library(ResourceSelection)###Hosmer-Lemeshow test

###Prepare/import data, label file as ‘data’. Include only columns that are to be included in the model.
###Place the dependent variable as the last column and label it as ‘Outcome’. Must be a binary factor
###Ensure that variables are labelled correctly (i.e. ordinal/categorical/continuous)

###Returns a dataframe, ‘dataC’ which contains only complete cases.
dataC<-na.omit(data)

###Multiple imputation (such as in the MICE package) may be useful if there is a large amount of missing
###data, however will introduce bias over-optimism of performance in Internal validation metrics and
###should be assessed on external data (or a hold-out set) unless models can be pooled using Rubin’s rules
###This is possible for logistic/linear/cox regression using package ‘rms’, but not for ML models at present.

###Confirms levels of the dependent variable
levels(dataC$Outcome)<- c(“No”, “Yes”)

###Returns a generic formula, ‘fm’ for the model
p<-Outcome
fm <- as.formula( paste( p, “.”, sep=“ ∼ “))

###Returns rules for model training and hyperparameter tuning.
###’logloss’ will result in 10 fold cross validation, repeated 5 times
###with the reported hyperparameters optimised for log-loss (which should optimise
###the probabilities of the predictions).
###’bootstrap’ will perform the 0.632 bootstrap with 1000 resamples.
###If used with fixed hyperparameters then this will internally validate the model

logloss=trainControl(method=“repeatedcv”,
              number=10,
              repeats=5,
              classProbs=TRUE,
              savePredictions=TRUE,
              summaryFunction = mnLogLoss)
bootstrap <- trainControl(method=“boot632”, number=1000,returnResamp = “all”,
                classProbs = TRUE, summaryFunction = twoClassSummary, savePredictions = TRUE)

####ELASTIC NET########
####Tunes hyperparameters and stores in ‘tuningmodel’. Can fix the alpha to a set level if desired.
tuningmodel<- train(fm, data=dataC, method = “glmnet”, trControl = logloss,metric = “logLoss”,
              tuneGrid = expand.grid(alpha = seq(0,1,by=0.1),lambda = seq(0.001,0.1,by = 0.001))) tuningmodel
###Trains the final model.
FinalEL<-train(fm, data=dataC, method = “glmnet”, trControl = bootstrap,metric = “ROC”,
               tuneGrid = expand.grid(alpha = tuningmodel$bestTune$alpha.,lambda =
tuningmodel$bestTune$lambda))

###Returns the apparent AUC from the final EL model
getTrainPerf(FinalEL)

###Trains Random Forest Model with hyperparameters according to ‘tgrid’
tgrid<-expand.grid(
 .mtry=2:10,
 .splitrule=c(“gini”,”extratrees”),
 .min.node.size=c(1,3,5)
)
RFModel<-train(fm, data=dataC, method=“ranger”, num.trees=1000, na.action=na.pass,replace=TRUE,
             trControl= logloss, tuneGrid=tgrid, metric =“ROC”)

###Trains the final Random Forest model
tgrid2<-expand.grid(
 .mtry=RFModel$bestTune$mtry,
 .splitrule=RFModel$bestTune$splitrule,
 .min.node.size=RFModel$bestTune$min.node.size
)

FinalRF<-train(fm, data=dataC, method=“ranger”, num.trees=1000, na.action=na.pass,replace=TRUE,
               trControl= bootstrap, tuneGrid=tgrid2, metric =“ROC”)

###Returns the apparent AUC from the final RF model
getTrainPerf(FinalRF)

####Trains XGB Model according to tuning parameters in ‘tune_grid’
####Note that this will take considerable time as it will train 25,200,000 models, consider adding parallel
processing if computer is multicore (add number of cores -1 after metric=“ROC” e.g. nthread =3 if 4 core
cpu)

tune_grid <- expand.grid(
 nrounds = seq(from = 100, to = 10000, by = 100),
 eta = c(0.025, 0.05, 0.1, 0.3),
 max_depth = c(2, 3, 4, 5, 6),
 gamma = c(0,0.05,0.1,0.5,0.7,0.9,1.0),
 colsample_bytree = c(0.4,0.6,0.8,1.0),
 min_child_weight = c(1,2,3),
 subsample = c(0.5,0.75,1.0)
)

xgb_tune <-train(fm,
                data=dataC,
                method=“xgbTree”,
                trControl=logloss,
                tuneGrid=tune_grid,
                verbose=T,
                metric=“ROC”)

####Returns Final XGB model

final_grid <- expand.grid(
 nrounds = xgb_tune$bestTune$nrounds,
 eta = xgb_tune$bestTune$eta,
 max_depth = xgb_tune$bestTune$max_depth,
 gamma = xgb_tune$bestTune$gamma,
 colsample_bytree = xgb_tune$bestTune$colsample_bytree,
 min_child_weight = xgb_tune$bestTune$min_child_weight,
 subsample = xgb_tune$bestTune$subsample
)

FinalXGB <-train(fm,
                  data=dataC,
                  method=“xgbTree”,
                  trControl=bootstrap,
                  tuneGrid=final_grid,
                  verbose=T,
                  metric=“ROC”)

###Returns the apparent AUC from the final XGB model
getTrainPerf(FinalXGB)

####Ensembles the Models into one model using linear blend

EnsList<-caretList(fm,data=dataC, trControl=logloss,
                tuneList=list(
                ranger=caretModelSpec(method=“ranger”, num.trees=1000,tuneGrid=tgrid2,
importance=“impurity”)),
                glmnet=caretModelSpec(method=“glmnet”,tuneGrid = expand.grid(alpha =
FinalEL$bestTune$alpha,lambda = FinalEL$bestTune$lambda)),
                xgbTree=caretModelSpec(method=“xgbTree”, tuneGrid=final_grid)
))

FinalEns<-caretEnsemble(
 EnsList,
 metric=“ROC”,
 trControl=trainControl(
  method=“repeatedCV”,
  number=10,
  repeats=5,
  classProbs=TRUE,
  savePredictions=TRUE,
  summaryFunction = mnLogLoss,
  verbose=TRUE
))

###Returns the apparent ROC and Calibration Chart
X<-predict(FinalEL, type=“prob”)
X1<-X[,2]
X2<-as.numeric(dataC[,o])-1
X3<-cbind2(X1,X2)

XR<-predict(FinalRF, type=“prob”)
X1R<-XR[,2]
X2R<-as.numeric(dataC[,o])-1
X3R<-cbind2(X1R,X2R)

XX<-predict(FinalXGB, type=“prob”)
X1X<-XX[,2]
X2X<-as.numeric(dataC[,o])-1
X3X<-cbind2(X1X,X2X)

XE<-predict(FinalEns,type=“prob”)
X1E<-XE
X2E<-as.numeric(dataC[,o])-1
X3E<-cbind2(X1E,X2E)

par(mfrow=c(2,2))

FinalModelROC <- plot.roc(X3[,2], X3[,1],
          main=“Elastic Net”,
          grid=TRUE, auc=TRUE,print.auc=TRUE, percent=TRUE,
          xlim=c(100, 0), ylim=c(0, 100),axis(1, at=c(100,0)),
          xlab=“Specificity (%)”, ylab=“Sensitivity (%)”,

                     ci=TRUE)
ciroccurve <- ci.se(FinalModelROC,specificities = seq(0, 100, 5), boot.n=1000)
plot(ciroccurve, type = “shape”, col = “lightgrey”)

RFModelROC <- plot.roc(X3R[,2], X3R[,1],
                   main=“RandomForest”,
                   grid=TRUE, auc=TRUE,print.auc=TRUE, percent=TRUE,
                   xlim=c(100, 0), ylim=c(0, 100),axis(1, at=c(100,0)),
                   xlab=“Specificity (%)”, ylab=“Sensitivity (%)”,
                   ci=TRUE)
ciroccurveR <- ci.se(RFModelROC,specificities = seq(0, 100, 5), boot.n=1000)
plot(ciroccurveR, type = “shape”, col = “lightgrey”)

XGBModelROC <- plot.roc(X3X[,2], X3X[,1],
            main=“XG Boost”,
            grid=TRUE, auc=TRUE,print.auc=TRUE, percent=TRUE,
            xlim=c(100, 0), ylim=c(0, 100),axis(1, at=c(100,0)),
            xlab=“Specificity (%)”, ylab=“Sensitivity (%)”,
            ci=TRUE)
ciroccurveX <- ci.se(XGBModelROC,specificities = seq(0, 100, 5), boot.n=1000)
plot(ciroccurveX, type = “shape”, col = “lightgrey”)

EnsModelROC <- plot.roc(X3E[,2], X3E[,1],
            main=“Ensemble”,
            grid=TRUE, auc=TRUE,print.auc=TRUE, percent=TRUE,
            xlim=c(100, 0), ylim=c(0, 100),axis(1, at=c(100,0)),
            xlab=“Specificity (%)”, ylab=“Sensitivity (%)”,
            ci=TRUE)
ciroccurveE <- ci.se(EnsModelROC,specificities = seq(0, 100, 5), boot.n=1000)
plot(ciroccurveE, type = “shape”, col = “lightgrey”)

{par(mfrow=c(2,2))
calibrate.plot(X3[,2],X3[,1], ylab=“Observed Probability”, xlab=“Predicted Probability”,main=“Elastic Net”)
calibrate.plot(X3R[,2],X3R[,1], ylab=“Observed Probability”, xlab=“Predicted Probability”, main=“Random
Forest”)
calibrate.plot(X3X[,2],X3X[,1], ylab=“Observed Probability”, xlab=“Predicted Probability”, main=“XG Boost”)
calibrate.plot(X3E[,2],X3E[,1], ylab=“Observed Probability”, xlab=“Predicted Probability”, main=“Ensemble”)
}

###Returns the Hosmer-Lemeshow test for the final model with 10 bins
HLFull<-hoslem.test(X3[,2],X3[,1],g=10)
HLFull
HLRFull<-hoslem.test(X3R[,2],X3R[,1],g=10)
HLRFull
HLXFull<-hoslem.test(X3X[,2],X3X[,1],g=10)
HLXFull
HLEFull<-hoslem.test(X3E[,2],X3E[,1],g=10)
HLEFull

###Internal Validation via bootstrapping - gives the final model ROC with 95%CI for 1000 bootstrap
samples
###(based on the ‘bootstrap’ trControl function)
par(mfrow=c(1,4))

BootELROC <- plot.roc(FinalEL$pred$obs, FinalELl$pred$Yes,
          main=“Bootstrap - Elastic Net”,
          grid=TRUE, auc=TRUE,print.auc=TRUE, percent=TRUE,
          xlim=c(100, 0), ylim=c(0, 100),axis(1, at=c(100,0)),
          xlab=“Specificity (%)”, ylab=“Sensitivity (%)”,
          ci=TRUE)

BootRFROC <- plot.roc(FinalRF$pred$obs, FinalRF$pred$Yes,
          main=“Bootstrap - Random Forest”,
          grid=TRUE, auc=TRUE,print.auc=TRUE, percent=TRUE,
          xlim=c(100, 0), ylim=c(0, 100),axis(1, at=c(100,0)),
          xlab=“Specificity (%)”, ylab=“Sensitivity (%)”,
          ci=TRUE)

BootXGBROC <- plot.roc(FinalXGB$pred$obs, FinalXGB$pred$Yes,
         main=“Bootstrap - XGB”,
         grid=TRUE, auc=TRUE,print.auc=TRUE, percent=TRUE,
         xlim=c(100, 0), ylim=c(0, 100),axis(1, at=c(100,0)),
         xlab=“Specificity (%)”, ylab=“Sensitivity (%)”,
         ci=TRUE)

BootEns <- plot.roc(FinalEns$ens_model$pred$obs, FinalEns$ens_model$pred$Yes,
                    main=“Bootstrap - XGB”,
                    grid=TRUE, auc=TRUE,print.auc=TRUE, percent=TRUE,
                    xlim=c(100, 0), ylim=c(0, 100),axis(1, at=c(100,0)),
                    xlab=“Specificity (%)”, ylab=“Sensitivity (%)”,
                    ci=TRUE)

###To perform internal-external validation - create datafiles with only and without
###each ‘centre’ to be tested. Then train models as above on each of the datasets with a centre missing,
###and use the ‘predict’ function on the dataset with only that centre to return results, example below.

Xa1<-predict(ELNa, newdata=data3, type=“prob”)
X1a1<-Xa1[,2]
X2a1<-as.numeric(data3$Outcome)-1
X3a1<-cbind2(X1a1,X2a1)

###Then combine the results for each centre weighted by the number of patients per centre. Note that if
predicting using new data, need to enter ‘newdata=‘ argument in the predict function. If using the training
data, do not put any argument, or put ‘data=‘ argument instead.

#####Probability Scaling using isotonic regression, used for non-probabilistic classifiers e.g. Tree based.
Isotonic regression function modified and derived from
https://www.analyticsvidhya.com/blog/2016/07/platt-scaling-isotonic-regression-minimize-logloss-error/
fit.isoreg <- function(iso, x0)
{
 o = iso$o
 if (is.null(o))
   o = 1:length(x)
 x = iso$x[o]
 y = iso$yf
 ind = cut(x0, breaks = x, labels = FALSE, include.lowest = TRUE)
 min.x <- min(x)
 max.x <- max(x)
 adjusted.knots <- iso$iKnots[c(1, which(iso$yf[iso$iKnots] > 0))]
 fits = sapply(seq(along = x0), function(i) {
   j = ind[i]

   if (is.na(j)) {
    if (x0[i] > max.x) j <- length(x)
    else if (x0[i] < min.x) j <- 1
   }
   upper.step.n <- min(which(adjusted.knots > j))
   upper.step <- adjusted.knots[upper.step.n]
   lower.step <- ifelse(upper.step.n==1, 1, adjusted.knots[upper.step.n -1] )
   denom <- x[upper.step] - x[lower.step]
   denom <- ifelse(denom == 0, 1, denom)
   val <- y[lower.step] + (y[upper.step] - y[lower.step]) * (x0[i] - x[lower.step]) / (denom)
   val <- ifelse(val > 1, max.x, val)
   val <- ifelse(val < 0, min.x, val)
   val <- ifelse(is.na(val), max.x, val)
   val
  })
  fits
}
Generate predictions, ideally on an external dataset (external_data) as described above using the predict function of caret. Then

XEN<-predict(FinalEns,newdata=external_data,type=“prob”)
X1EN<-XEN
X2EN<-as.numeric(external_data[,o])-1
########Below returns data before adjustment
X3EN<-cbind2(X1EN,X2EN)
########Below returns data after adjustment.
iso.model<-isoreg(X3EN[,1],X3EM[,2])
X1ENs<-fit.isoreg(iso.model,X1EN)
X3ENa<-cbind2(((X1ENs*0.9999999)+0.000000001),X2EN)
####Calibration can then be reassessed as above. For this method of calibration chart, must make sure
that the values are not exactly 0 or 1, hence the multiplication/addition above.

#######Final Model Variable Importance
varImp<-varImp(FinalEns)

citation(‘caret’)
citation(‘pROC’)
citation(‘ResourceSelection’)
citation(‘caretEnsemble’)
citation(‘gbm’)

#### End

#####To use the models provided at https://uoscancer.shinyapps.io/EROC/

#####**Prepare test data to match model**;
#####Column names (case sensitive): Gender, Age, Site, NAResponder, pT, VascInv, R0, Grade, NPosLN,
Nodes_Examined, EarlyRec.
#####NAResponder should be TRG 1-2 or TRG3-5 and Grade should be Good, Moderate, Poor/Anaplastic
#####Set Gender, Site, NAResponder, VascInv, R0, EarlyRec to factors and Age, pT, Grade, NPosLN and
Nodes_Examined to numeric
#####Use mapvalues (case sensitive) so that gender is “Male”/”Female”, Site is “GOJ”/”Oesophagus”, NAResponder is “TRG1-2”/”TRG3-5”, VascInv is “No”/”Yes”, R0 is “R0”/”R1”, EarlyRec is “No”/”Yes”
#####Load model and isotonic model into workspace

####Run isotonic fit function (fit.isoreg, as above)

####Obtain predictions using the model calibrated using isotonic regression
XE<-predict(Ensemble, newdata=data, type=“prob”)
X1E<-1-XE
X1Ea<-(fit.isoreg(iso.model,X1E)*0.99999+0.0000000001)
X2E<-as.numeric(data$EarlyRec)-1
X3E<-cbind2(X1Ea,X2E)

####Use predictions as above
~~~

## Appendix

### Oesophageal Cancer Clinical and Molecular Stratification (OCCAMS) Consortium Members List

Ayesha Noorani^1^, Rachael Fels Elliott^1^, Paul A.W. Edwards^1,2^, Nicola Grehan^1^, Barbara Nutzinger^1^, Jason Crawte^1^, Hamza Chettouh^1^, Gianmarco Contino^1^, Xiaodun Li^1^, Eleanor Gregson^1^, Sebastian Zeki^1^, Rachel de la Rue^1^, Shalini Malhotra^1,3^, Simon Tavaré^2^, Andy G. Lynch^2^, Mike L. Smith^2^, Jim Davies^5^, Charles Crichton^5^, Nick Carroll^6^, Peter Safranek^6^, Andrew Hindmarsh^6^, Vijayendran Sujendran^6^, Stephen J. Hayes^7,14^, Yeng Ang^7,8,29^, Shaun R. Preston^9^, Sarah Oakes^9^, Izhar Bagwan^9^, Vicki Save^10^, Richard J.E. Skipworth^10^, Ted R. Hupp^10^, J. Robert O’Neill^10,23^, Olga Tucker^11,33^, Andrew Beggs^11,28^, Philippe Taniere^11^, Sonia Puig^11^, Timothy J. Underwood^12,13^, Fergus Noble^12^, Jack Owsley^12^, Hugh Barr^15^, Neil Shepherd^15^, Oliver Old^15^, Jesper Lagergren^16,25^, James Gossage^16,24^, Andrew Davies^16,24,^ Fuju Chang^16,24^, Janine Zylstra^16,24^, Vicky Goh^24^, Francesca D. Ciccarelli^24^, Grant Sanders^17^, Richard Berrisford^17^, Catherine Harden^17^, David Bunting^17^, Mike Lewis^18^, Ed Cheong^18^, Bhaskar Kumar^18^, Simon L. Parsons^19^, Irshad Soomro^19^, Philip Kaye^19^, John Saunders^19^, Laurence Lovat^20^, Rehan Haidry^20^, Victor Eneh^20^, Laszlo Igali^21^, Michael Scott^22^, Shamila Sothi^26^, Sari Suortamo^26^, Suzy Lishman^27^, George B. Hanna^31^, Christopher J. Peters^31^, Anna Grabowska^32^

^1^Medical Research Council Cancer Unit, Hutchison/Medical Research Council Research Centre, University of Cambridge, Cambridge, UK; ^2^Cancer Research UK Cambridge Institute, University of Cambridge, Cambridge, UK; ^3^Department of Histopathology, Addenbrooke’s Hospital, Cambridge, UK; ^4^Oxford ComLab, University of Oxford, UK, OX1 2JD; ^5^Department of Computer Science, University of Oxford, UK, OX1 3QD; ^6^Cambridge University Hospitals NHS Foundation Trust, Cambridge, UK, CB2 0QQ; ^7^Salford Royal NHS Foundation Trust, Salford, UK, M6 8HD; ^8^Wigan and Leigh NHS Foundation Trust, Wigan, Manchester, UK, WN1 2NN; ^9^Royal Surrey County Hospital NHS Foundation Trust, Guildford, UK, GU2 7XX; ^10^Edinburgh Royal Infirmary, Edinburgh, UK, EH16 4SA; ^11^University Hospitals Birmingham NHS Foundation Trust, Birmingham, UK, B15 2GW; ^12^University Hospital Southampton NHS Foundation Trust, Southampton, UK, SO16 6YD; ^13^Cancer Sciences Division, University of Southampton, Southampton, UK, SO17 1BJ; ^14^Faculty of Medical and Human Sciences, University of Manchester, UK, M13 9PL; ^15^Gloucester Royal Hospital, Gloucester, UK, GL1 3NN; ^16^St Thomas’s Hospital, London, UK, SE1 7EH; ^17^Plymouth Hospitals NHS Trust, Plymouth, UK, PL6 8DH; ^18^Norfolk and Norwich University Hospital NHS Foundation Trust, Norwich, UK, NR4 7UY; ^19^Nottingham University Hospitals NHS Trust, Nottingham, UK, NG7 2UH; ^20^University College London, London, UK, WC1E 6BT; ^21^Norfolk and Waveney Cellular Pathology Network, Norwich, UK, NR4 7UY; ^22^Wythenshawe Hospital, Manchester, UK, M23 9LT; ^23^Edinburgh University, Edinburgh, UK, EH8 9YL; ^24^King’s College London, London, UK, WC2R 2LS; ^25^Karolinska Institutet, Stockholm, Sweden, SE-171 77; ^26^University Hospitals Coventry and Warwickshire NHS, Trust, Coventry, UK, CV2 2DX; ^27^Peterborough Hospitals NHS Trust, Peterborough City Hospital, Peterborough, UK, PE3 9GZ; ^28^Institute of Cancer and Genomic sciences, University of Birmingham, B15 2TT; ^29^GI science centre, University of Manchester, UK, M13 9PL; ^30^Queen’s Medical Centre, University of Nottingham, Nottingham, UK, NG7 2UH; ^31^Imperial College NHS Trust, Imperial College London, UK, W2 1NY; ^32^Queen’s Medical Centre, University of Nottingham, Nottingham, UK; ^33^Heart of England NHS Foundation Trust, Birmingham, UK, B9 5SS.

